# Association of social isolation, loneliness, and genetic risk with incidence of dementia: UK Biobank cohort study

**DOI:** 10.1101/2020.02.25.20027177

**Authors:** Marko Elovainio, Jari Lahti, Matti Pirinen, Laura Pulkki-Råback, Anni Malmberg, Jari Lipsanen, Marianna Virtanen, Mika Kivimäki, Christian Hakulinen

## Abstract

**Objective:** To examine the associations of social isolation and loneliness with incident dementia by level of genetic risk.

**Design:** Prospective population-based cohort study.

**Setting and participants:** 155 074 men and women (mean age 64.1, SD 2.9 years) from the UK Biobank Study, recruited between 2006 and 2010.

**Main exposures:** Self-reported social isolation and loneliness, and polygenic risk score for Alzheimer’s disease with low (lowest quintile), intermediate (quintiles 2 to 4), and high (highest quintile) risk categories.

**Main outcome:** Incident all-cause dementia ascertained using electronic health records.

**Results:** Overall, 8.6% of participants reported that they were socially isolated and 5.5% were lonely. During a mean follow-up of 8.8 years (1.36 million person-years), 1444 (0.9% of the total sample) were diagnosed with dementia. Social isolation, but not loneliness, was associated with increased risk of dementia (hazard ratio 1.62, 95% confidence interval 1.38 to 1.90). Of the participants who were socially isolated and had high genetic risk, 4.2% (2.9% to 5.5%) were estimated to develop dementia compared with 3.1% (2.7% to 3.5%) in participants who were not socially isolated but had high genetic risk. The corresponding estimated incidence in the socially isolated and not isolated were 3.9% (3.1% to 4.6%) and 2.5% (2.2% to 2.6%) in participants with intermediate genetic risk.

**Conclusion:** Socially isolated individuals are at increased risk of dementia at all levels of genetic risk.

**What is already known on this topic:**

- Social isolation and loneliness have been associated with increased risk of dementia
- It is not known whether this risk is modified or confounded by genetic risk of dementia

**What this study adds:**

- This is the first study to show that social isolation is associated with increased risk of dementia across the spectrum of genetic risk
- Loneliness, although considered as a significant risk for multiple health problems, seems to be associated with dementia only when combined with high genetic risk

---

The rapidly rising numbers of people with dementia ^1^ is a significant health policy and health service concern in many high-income countries. Although considerable share of the dementia risk is due to genetic factors ^2-4^, major efforts have been directed towards the identification of potentially modifiable risk factors that could prevent or delay the onset of dementia ^5^. Higher levels of social support have been suggested to protect from dementia ^6^, with both social isolation and feelings of loneliness being associated with increased risk of dementia ^7-10^, although mixed findings have reported between loneliness and dementia risk ^11 12^. However, it remains unclear whether there is an interplay between genetic and social isolation and loneliness (*i*.*e*. whether the association of social isolation and loneliness with dementia is evident only at high or low levels of genetic risk) or whether the associations of genetic factors and social support with dementia are independent and additive.

The polygenic risk score (PRS) for Alzheimer’s disease, describing the polygenic burden captured by the most recent genome-wide studies ^13^, allows to estimate the size of the genetic risk and the extent to which the associations of social isolation and loneliness with dementia are modified by genetic risk. In the present study, we used data from UK Biobank study to examine whether genetic risk may intensify and attenuate the associations of social isolation and loneliness with the risk of dementia. In addition to estimating relative risk, we will provide estimates of absolute risk ^14^, as they are important information for risk communication and clinical risk prediction ^15^.

## METHODS

### Study design and participants

In this analysis of the UK Biobank study, we used baseline data and obtained information of incident dementia at follow-up via linked electronic health records ^16^. UK National Health Service (NHS) registers maintain records of all individuals legally registered as residents in the United Kingdom. In the UK Biobank study, these records were used to invite around 9.2 million individuals aged 40–69 years living within a sensible travelling distance of the 22 assessment centres across Great Britain 2007–2010 ^16^. At the study baseline, participants completed multiple touchscreen computer-based questionnaires followed by a face-to-face interview with trained research staff. Physical measures were also taken. Details of these assessments and variables are publicly available from the UK Biobank website: http://biobank.ctsu.ox.ac.uk/crystal/.

In total, 502,656 individuals were recruited (5.4% of the eligible population). Of those, individuals that were 60 year or older and had complete data on social isolation, loneliness, dementia and genetic data were included in the present analysis (N = 147 614 – 152 070). We also repeated the analyses using imputed data in those with missing information on social isolation, loneliness or other explanatory variables but had information on genetic risk score (N = 155 070). This study was conducted under generic approval from the NHS National Research Ethics Service (17th June 2011, Ref 11/NW/0382). Participants provided electronic consent for the baseline assessments and register linkage.

### Ascertainment of incident dementia

Dementia was ascertained using hospital inpatient records which contains data on admissions and diagnoses from the Hospital Episode Statistics for England, Scottish Morbidity Record data for Scotland, and the Patient Episode Database for Wales. Additional cases were detected through linkage to death register data provided by the National Health Service Digital for England and Wales and the Information and Statistics Division for Scotland. Diagnoses were recorded using the International Classification of Diseases (ICD) coding system. Participants with dementia were identified as having a primary/secondary diagnosis (hospital records) or underlying/contributory cause of death (death register) using ICD-9 and ICD-10 codes for Alzheimer disease and other dementia classifications (see the online supplement for details).

### Measurement of social isolation and loneliness

Social isolation and loneliness were measured using the same scale as in our two previous UK Biobank studies ^17 18^. *Social isolation* scale was defined using the following three questions: (a) “Including yourself, how many people are living together in your household? Include those who usually live in the house such as students living away from home during term, partners in the armed forces or professions such as pilots” (1 point for living alone) (b) “How often do you visit friends or family or have them visit you?” (1 point for friends and family visits less than once a month), and (c) “Which of the following [leisure/social activities] do you attend once a week or more often? You can select more than one”, (1 point for no participation in social activities at least weekly). This resulted in scale with a range from 0 to 3, where an individual was defined as socially isolated if he/she had two or more of those points and those who scored 0 or 1 were classified as not isolated. Other studies in the UK have used similar measures ^18^.

*Loneliness* scale was constructed from two questions: “Do you often feel lonely?” (no = 0, yes=1) and ““How often are you able to confide in someone close to you?”(0 = almost daily-once every few months 1= once every few months to never or almost never). An individual was defined as lonely if he/she responded positively to both questions (score 2) and not lonely if he or she responded negatively to one or both of the questions (score 0 -1). Similar questions have been used in longer loneliness scales, such as the Revised UCLA Loneliness Scale ^19^.

### Polygenic risk score of dementia

From the genotyped UK Biobank samples, we included 155,070 unrelated white British participants after removal of participants based on heterozygosity and missingness of outliers, sex chromosome aneuploidies and mismatches, withdrawals, and those that UK Biobank had excluded from the relatedness calculations. The genotypes were imputed against Haplotype Reference Consortium and UK10K haplotype resources containing ∼96M variants ^13^. We calculated polygenic risk scores (PRS) for Alzheimer’s disease (AD) based on a genome-wide association study by Kunkle and others (2019) with 35,274 AD cases and 59,163 controls that do not overlap with UK Biobank samples (for details see the online supplement). We used Plink 1.9 ^20^ for the genotype QC and clumping. The following parameters were used for the clumping of the genotype data: p-value threshold 0.5, LD threshold (r^2^) 0.5, and clumping window width of 250 kilobases. Prior to clumping we excluded all SNPs with MAF < 0.001, genotyping rate < 0.1, Hardy-Weinberg equilibrium p-value < 1e-6 and missingness per person >0.1. We used PRSice 2.2.8 ^21^ for calculating the PRS with the genotype QC settings that have been recommended by the software developers ^22^. In the main analyses, we applied a p-value threshold of 0.5, which resulted in including 626,623 SNPs in the PRS. This threshold was chosen as previous work has reported that it provided an optimal set of variants for predicting dementia and AD ^23 24^. While this set is likely to include a number of variants which are not associated with AD, it also includes a number of variants that at present do not have sufficient statistical evidence to meet the criteria for being genome-wide significant (i.e. P-value < 5×10-8) but are expected to be associated in future larger studies. The univariate associations between genetic risks score with 10 different cut-off points and incident dementia is reported in the online supplement (SFigure 1).

The polygenic risk scores were then z-standardized to have mean 0 and variance 1, divided into quintiles and categorized as low (lowest quintile), intermediate (quintiles 2 to 4) and high (highest quintile).

### Assessment of potential explanatory factors

Following information was used in the current study: sex, age in years, socioeconomic factors (educational attainment and Townsend deprivation index, which is an area-level composite measure of deprivation based on unemployment, non-home ownership, non-car ownership, and household overcrowding), chronic diseases (diabetes, cardiovascular disease, cancer, and other long-standing illness, disability or infirmity), cigarette smoking (smoker [yes/no]; ex-smoker[yes/no]), physical activity (moderate and vigorous physical activity), alcohol intake frequency (Three or four times a week or more vs. once or twice a week or less), and the frequency of depressed mood in the past 2 weeks (Patient Health Questionnaire; ^25^).

### Statistical analyses

Study participants were followed from the study baseline (2006-2010) for incident dementia until the date of first dementia diagnosis, death, or to the end of the follow-up, whichever came first. The associations of social isolation, loneliness and polygenic risk score with incident dementia were examined using Cox proportional hazard regression models where age was used as a time scale. Results from these analyses were reported as hazard ratios (relative risk) and their 95% confidence intervals and the models were adjusted for age, sex, and 10 first principal components of genetic structure from UK Biobank to control for possible population stratification, and additionally for education, social deprivation index, having long term illness, physical activity, smoking status, alcohol consumption, and depressive symptoms. In these analyses, PRS was used both as a categorical and as a continuous variable. Cumulative incidence (absolute risk) of dementia associated with categories of social isolation, loneliness and genetic risk was estimated using competing-risks regression ^26-28^, with death being treated as competing event.

Missing data on social isolation, loneliness and all explanatory factors were imputed using multiple imputation by chained equations to generate five imputed datasets. Imputation model included age, sex, social isolation, loneliness, all covariates, the Nelson-Aalen estimate of cumulative hazard, and survival status ^29^. Cox proportional hazards models were fitted within each imputed dataset and combined using Rubin’s rules. Frequencies of complete and imputed variables are reported in the online supplement table 3. P-values were 2-sided with statistical significance set at less than .05. All analyses were performed using Stata (15.1) and R (3.6.2).

### Patient involvement

These results are based on existing data. We were not involved in the recruitment of the participants. As far as we know, no patients were engaged in designing the present research question or the outcome measures. They were also not involved in developing plans for recruitment, design, or implementation of the study, and were not asked to advise on interpretation or writing up of results. Results from UK Biobank are disseminated to study participants via the study website and social media outlets.

## RESULTS

Descriptive statistics of the study participants are shown in **Table 1**. Genetic risk score data were available for 155 070 participants (51.9% women; mean age 64.1 years). Overall, 8.6% of participants (N = 13103) were classified as socially isolated and 5.5% were lonely (N = 8102). During a total of 1.36 million person-years (mean follow-up time 8.8 years), 1444 participants (0.9% of the total sample) were diagnosed with all-cause dementia.

**Table 1.** Baseline Characteristics of Participants according to diagnosed Dementia at follow-up Dementia.

|  |  | Dementia |  | p-value |
| --- | --- | --- | --- | --- |
| Variables |  | No | Yes |  |
| Age at baseline | Mean (SD) | 64.1 (2.8) | 65.8 (2.7) | <0.001 |
| Sex | Female | 79821 (52.0) | 631 (43.7) | <0.001 |
|  | Male | 73805 (48.0) | 813 (56.3) |  |
| Education | Lower | 40578 (26.7) | 536 (38.2) | <0.001 |
|  | Intermediate | 71839 (47.3) | 606 (43.2) |  |
|  | Higher | 39307 (25.9) | 261 (18.6) |  |
| Long term illness | No | 57738 (38.7) | 319 (23.3) | <0.001 |
|  | Yes | 91266 (61.3) | 1053 (76.7) |  |
| Physical activity | Low | 45963 (30.7) | 479 (34.9) | 0.001 |
|  | High | 103938 (69.3) | 893 (65.1) |  |
| Current smoker | No | 140646 (92.0) | 1281 (89.4) | <0.001 |
|  | Yes | 12265 (8.0) | 152 (10.6) |  |
| Alcohol consumption | Lower | 81242 (52.9) | 866 (60.1) | <0.001 |
|  | Higher | 72283 (47.1) | 575 (39.9) |  |
| Depressive symptoms | Low | 121508 (82.5) | 1014 (75.8) | <0.001 |
|  | Low-medium | 21350 (14.5) | 245 (18.3) |  |
|  | High_medium | 2788 (1.9) | 42 (3.1) |  |
|  | High | 1639 (1.1) | 37 (2.8) |  |
| Townsend deprivation index | Mean (SD) | -1.7 (2.8) | -1.1 (3.3) | <0.001 |
| Socially isolated (no / yes) | No | 138408 (91.5) | 1208 (87.3) | <0.001 |
|  | Yes | 12928 (8.5) | 175 (12.7) |  |
| Feelling lonely (no / yes) | No | 138255 (94.5) | 1253 (92.5) | 0.001 |
|  | Yes | 8000 (5.5) | 102 (7.5) |  |
| Genetic dementia risk | Low | 30834 (20.1) | 180 (12.5) | <0.001 |
|  | Intermediate | 92148 (60.0) | 895 (62.0) |  |
|  | High | 30644 (19.9) | 369 (25.5) |  |

As expected, a higher PRS for AD was associated with an increased risk of dementia. Using continuous PRS, the hazard ratio per 1SD increase in the score was 1.27 (95% CI 1.21 to 1.34) in an analysis adjusted for age, sex and 10 principal components. The associations between genetic risk categories (low, intermediate, and high) with incidence of dementia shown in **Table 2**. In comparison to the participants in the low category, the hazard ratio of incident dementia was 1.56 (95% CI 1.31 – 1.87) in participants with intermediate risk and 1.89 (95% CI 1.55 – 2.31) in those with high genetic risk in the fully adjusted model.

**Table 2.** Risk of Incident Dementia According to Genetic Risk, Social Isolation and Loneliness Categories.

|  | **Model 1** |  | **Model 2** |  |
| --- | --- | --- | --- | --- |
| *Genetic risk* | *Hazard Ratio*<br>*95 % CI* | *P-Value* | *Hazard Ratio*<br>*95 % CI* | *P-Value* |
| Intermediate vs. low | 1.66<br>(1.41 – 1.94) | **<0.001** | 1.56<br>(1.31 – 1.87) | **<0.001** |
| High vs. low | 2.06<br>(1.72 – 2.46) | **<0.001** | 1.89<br>(1.55 – 2.31) | **<0.001** |
| Observations | 155074 |  | 132628 |  |
| R<sup>2</sup> Nagelkerke | 0.345 |  | 0.461 |  |
|  | *Hazard ratio*<br>*95 % CI* | *P-Value* | *Hazard ratio*<br>*95 % CI* | *P-Value* |
| Separate analyses |  |  |  |  |
| Isolated vs no isolated | 1.62<br>(1.38 – 1.90) | **<0.001** | 1.33<br>(1.12 – 1.60) | **0.002** |
| Observations | 152723 |  | 137903 |  |
| R<sup>2</sup> Nagelkerke | 0.320 |  | 0.455 |  |
| Lonely vs not lonely | 1.47<br>(1.20 – 1.80) | **<0.001** | 1.03<br>(0.81 – 1.30) | 0.820 |
| Observations | 147614 |  | 133893 |  |
| R<sup>2</sup> Nagelkerke | 0.316 |  | 0.463 |  |
| Combined analysis |  |  |  |  |
| Lonely | 1.28<br>(1.03 – 1.59) | **0.025** | 0.95<br>(0.74 – 1.22) | 0.686 |
| Isolated | 1.58<br>(1.34 – 1.86) | **<0.001** | 1.33<br>(1.10 – 1.60) | **0.003** |
| Observations | 145663 |  | 132628 |  |
| R<sup>2</sup> Nagelkerke | 0.322 |  | 0.461 |  |

Model 1. Adjusted for age and sex
Model 2. Adjusted for age, sex, education, social deprivation, depressive symptoms, health behaviors, genetic risk score, and 10 principal components

Social isolation was associated with increased risk of dementia (HR adjusted for age and sex = 1.62, 95% CI 1.38 – 1.90). The associations attenuated but remained statistically significant after adjusting for additional covariates including socio-demographics, health-related factors and genetic risk score and principal components (HR = 1.33, 95% CI 1.12 – 1.60). Loneliness was also associated with higher risk of dementia (HR = 1.47, 95% CI 1.20 – 1.80), but this association was lost when adjusted for socio-demographics, health-related factors, PRS and principal components (HR = 1.03, 95% CI 0.81 – 1.30). Both social isolation (HR = 1.58, 95% CI 1.34 – 1.86) and loneliness (HR = 1.28, 95% CI 1.03 – 1.59) were associated with incident dementia when added simultaneously into the model but only the association between social isolation and dementia was robust to adjusting for additional covariates (HR = 1.33, 95 % CI 1.10 – 1.60). (**Table 2**)

When the interplay between genetic risk and social isolation was assessed using combined categories, there was a monotonic association of increasing genetic risk and social isolation with increasing dementia risk. In the fully adjusted models, compared to those with a low genetic risk and no social isolation, the isolated participants with a low genetic risk had a hazard ratio of 1.42 (95% CI, 0.88-2.27). The corresponding hazard ratios were 1.57, (95% CI 1.30 - 1.89) and 1.99, 95% CI 1.61 - 2.45) for those with intermediate or high genetic risk and no social isolation, and 2.16, (95% CI 1.65 – 2.83) and 2.37, 95% CI 1.62 – 3.46) for those who were socially isolated and had intermediate or high genetic risk (**Figure 1**). The results for loneliness were less consistent, although the risk of dementia was greater in lonely participants at low or at high levels of genetic risk, when compared with those who reported no loneliness. In the high genetic risk group, for example, the hazard ratios were 1.93 (95% CI 1.56 - 2.37) in low and 2.20 (95% CI 1.39-3.47) in high loneliness group (**Figure 2**).

**Figure 1.**
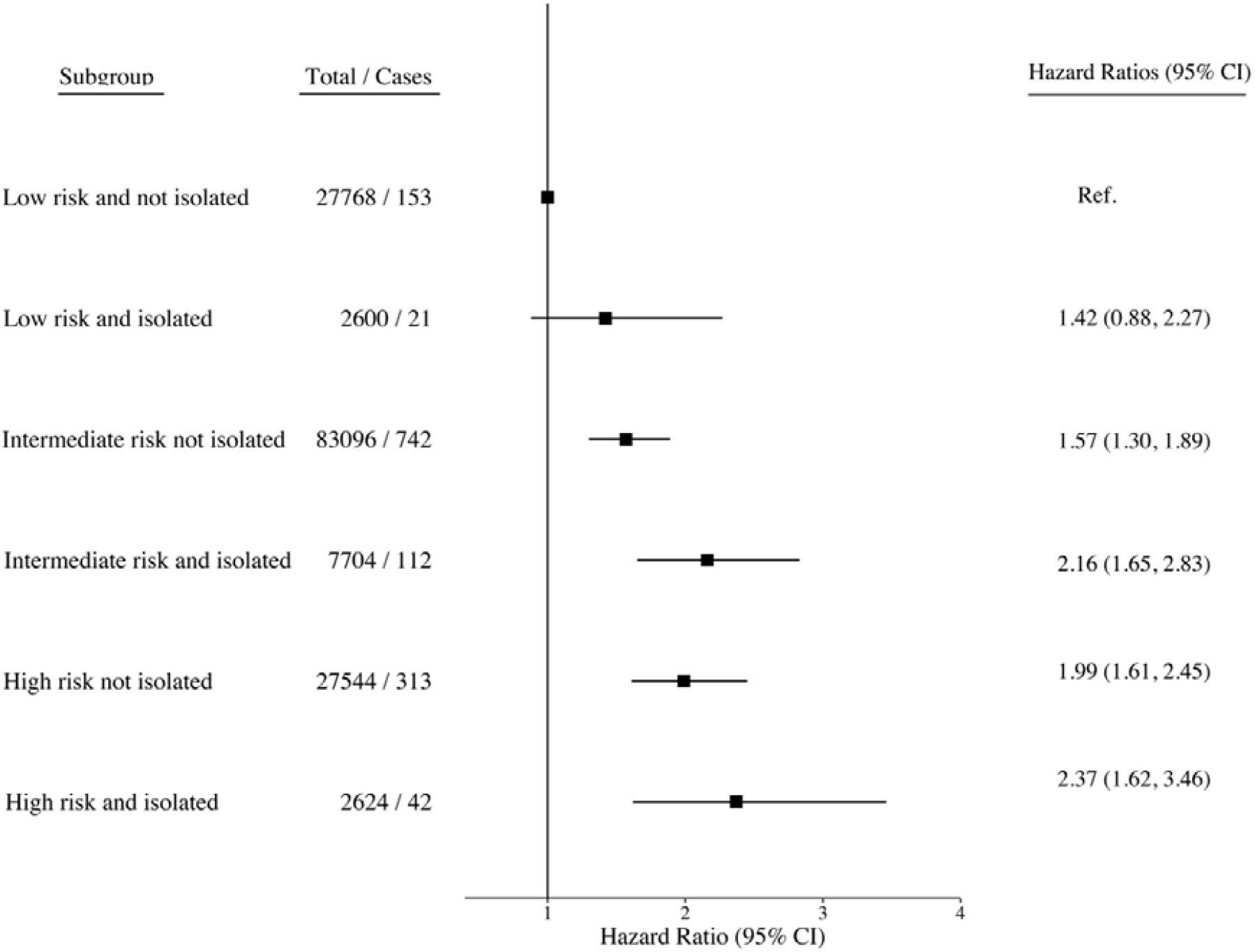
Associations of combined genetic risk and social isolation with incident dementia risk.

**Figure 2.**
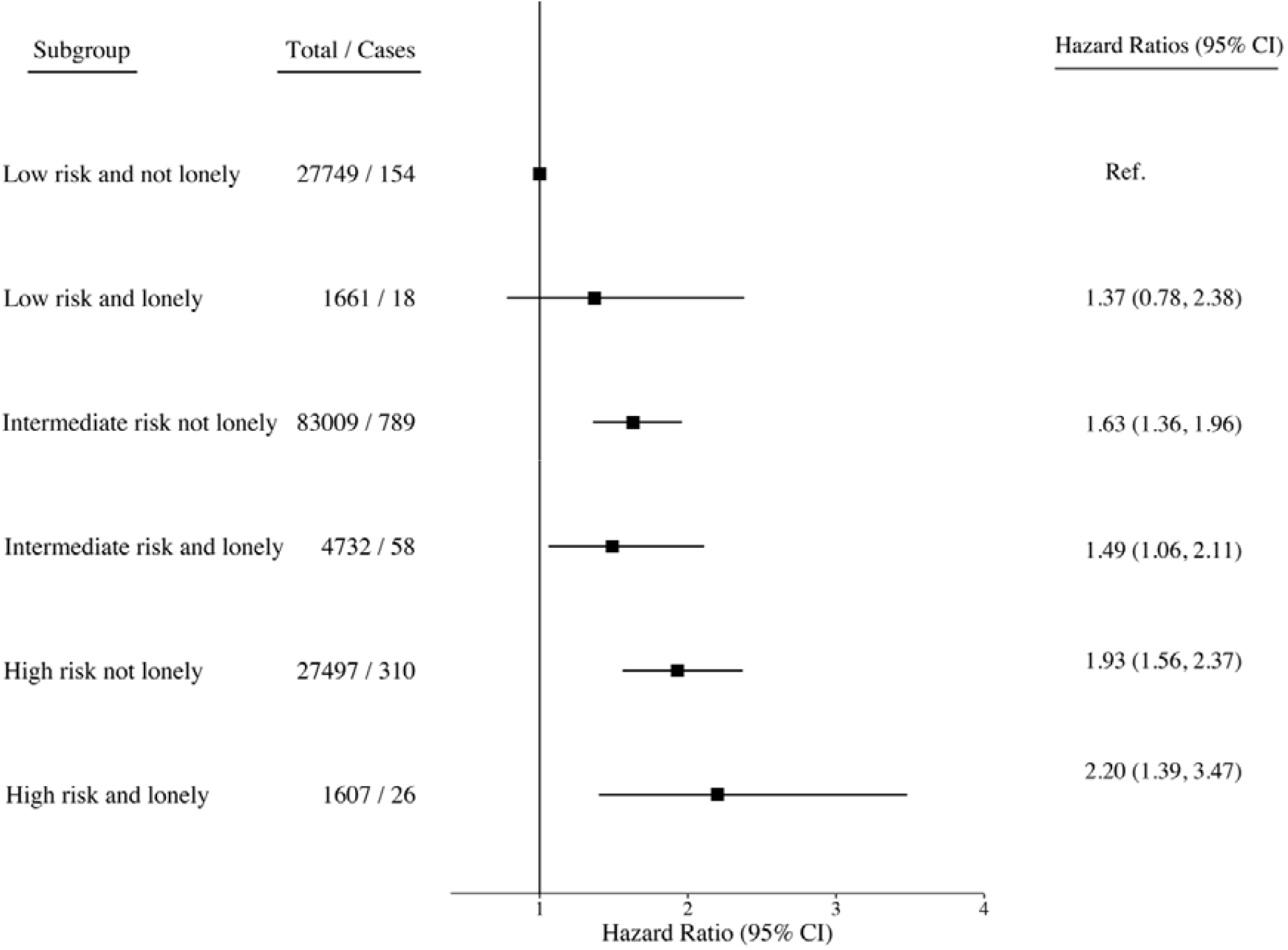
Associations of combined genetic risk and loneliness with incident dementia risk.

In terms of absolute risk (cumulative incidence), of those who were socially isolated and had high genetic risk, 4.2% (2.9% to 5.5%) were estimated to developed dementia compared with 3.1% (2.7% to 3.5%) of those who were not socially isolated but had high genetic risk (**Figure 3**). The corresponding absolute risk estimates in the socially isolated and not isolated were 3.9% (3.1% to 4.6%) and 2.4% (2.2% to 2.6%) in participants with intermediate genetic risk and 2.2% (1.2% to 3.1%) and 1.5% (1.2% to 1.7%) in those with low genetic risk.

**Figure 3.**
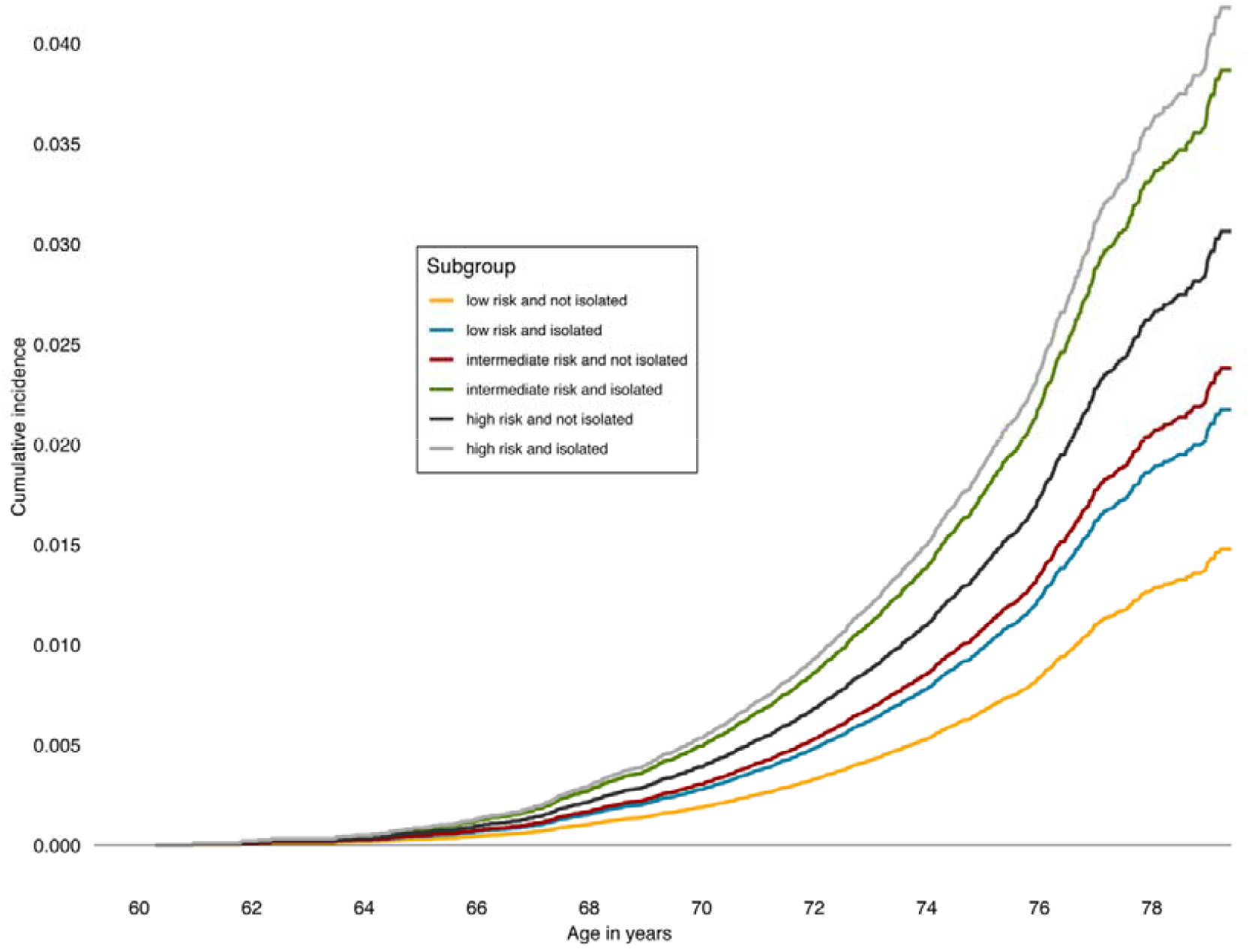
Cumulative incidence of dementia in combined genetic risk and social isolation groups.

As sensitivity analyses, we repeated all the main analyses with Alzheimer’s disease as the outcome and using imputed data sets (Supplement SFigures 2 – 3). The results did not materially change.

## DISCUSSION

In this UK Biobank study of 155 074 men and women, social isolation was associated with increased risk of all-cause dementia and Alzheimer’s disease at all levels of genetic risk of Alzheimer’s disease. The incidence of dementia was estimated to reach over 4% in isolated high-genetic risk individuals compared to approximately 3% in non-isolated individuals with similar genetic risk, the difference between these groups being over 1% also among those with intermediate and low genetic risk. This means that among individuals with similar genetic risk for dementia, those who are socially isolated are more likely to have incidence of the disease, suggesting an effect by social isolation over and above that of genetic risk. The association between loneliness and dementia was attributable to other dementia risk factors, such as health behaviours and depressive symptoms.

To the best of our knowledge, this is the first study examining the joint associations of aspects of social support and genetic risk with the incidence of dementia. The relative risk of dementia across the genetic risk categories was at the same magnitude as in a previous UK Biobank study ^30^ that used data from an older GWAS ^31^. Our findings also support other studies - most of which with follow-ups from 5 to 11 years – showing an association of social isolation with increased risk of dementia ^7 10 12^. A 28-year follow-up of 10,000 Whitehall II study participants found that less frequent social contacts at ages 50, 60 and 70 were associated with approximately 10% higher dementia risk, independent of socio-economic and other lifestyle factors ^32^. While previous studies have produced mixed findings on whether loneliness is associated with increased risk of dementia or not ^11 12^, our findings show that the association between loneliness and dementia is mostly likely explained by other factors and present only at high levels of genetic risk.

Our results should be interpreted in a context of disease aetiology. Dementia is characterised by a 10-20-year preclinical or prodromal stage during which changes in biomarkers and cognitive abilities increasingly occur ^33^. With a follow-up less than 10 years, it is likely that we assessed social isolation for dementia cases during this preclinical period. This could result to reverse causality, i.e., increased prevalence of social isolation during the 8-year period could have resulted from preclinical changes in social activity leading to a spurious association between social isolation and dementia.

Several mechanisms through which social isolation may causally affect dementia risk have been proposed. Social isolation and loneliness have been suggested to increase stress reactivity which is associated with prolonged activation of the hypothalamic-pituitary-adrenal axis (HPA) and the sympatho-adrenal system ^34^. This process may further lead to sleep deprivation, dysregulation of the immune system, and even increased levels of oxidative stress ^35-40^, all potentially harmful for cognitive health. It has also been shown that socially isolated and lonely individuals more often engage in health-damaging behaviors ^18 41^, which may affect cognition either directly via biophysiological mechanisms or increased incidence of cardiometabolic diseases which accelerate neurodegeneration ^42-45^. Socially isolated or lonely individuals are also at an increased risk of depression ^46^, a potential risk factor for cognitive decline and dementia ^47^. Participation in social activities and social interaction stimulates neural plasticity by building and maintaining cognitive reserve ^48 49^. Poor cognitive reserve is a further pathway through which social isolation and loneliness could increase dementia risk ^50^. Fewer social contacts with reduced exercising of memory and language adversely affect cognitive reserve, thereby accelerating dementia onset ^50^. Cognitive ability was not assessed in the present study and a small share of the found association between social isolation and subsequent dementia risk may be attributable to lower initial cognitive reserve.

### Strengths and limitations

The major strengths of the current study include the large sample size of UK Biobank participants, which enabled us to study the combination of genetic risk, social isolation, and loneliness in detail. In addition, we used the largest genome-wide association study of dementia to date to derive the genetic risk for AD ^2^.

There are also some important limitations. Although our analyses were adjusted for multiple potential sources of bias, the possibility of unmeasured confounding and reverse causation cannot be ruled out. Both frequency of social contacts and loneliness were self-reported and measured by relatively short and crude measures. As we were able to cover the genetic risk for AD – not all-cause dementias – based on the Kunkle et al ^2^, we may have missed some of the genetic variance related to non-AD dementias. Dementia cases were derived from medical records or death registers, and thus some cases might have been missed. However, good agreement of dementia case determination with primary care record data has been shown ^51^. This sample was restricted to volunteers of European ancestry aged 60 to 73 years at baseline and, therefore, further research is needed to ensure generalizability of our findings. As the mean age of participants was only 72 years at the end of the follow-up period, the incidence of dementia remained low. As noted previously the response rate of the UK Biobank study survey was very low, 5.5%, and UK Biobank is not representative of the sampling population ^52^. However, many etiological findings from UK Biobank appear to be generalisable to England and Scotland ^53^.

### Conclusions

The present findings suggest an association between social isolation and increased risk of dementia across the spectrum of genetic risk. Further research is needed to determine the extent social isolation is a modifiable risk factor rather than a part of the dementia prodrome

## Data Availability

The genetic and phenotypic UK Biobank data are available on application to the UK Biobank (www.ukbiobank.ac.uk/). Present study was conducted using the UK Biobank Resource under Application 14801. Summary statistics from the meta-analysis of genome wide association studies in dementia are available from https://www.niagads.org/datasets/ng00075

## Acknowledgments

We thank the International Genomics of Alzheimer’s Project (IGAP) for providing summary results data for these analyses. The investigators within IGAP contributed to the design and implementation of IGAP and/or provided data but did not participate in analysis or writing of this report. IGAP was made possible by the generous participation of the control subjects, the patients, and their families. The i–Select chips was funded by the French National Foundation on Alzheimer’s disease and related disorders. EADI was supported by the LABEX (laboratory of excellence program investment for the future) DISTALZ grant, Inserm, Institut Pasteur de Lille, Université de Lille 2 and the Lille University Hospital. GERAD/PERADES was supported by the Medical Research Council (Grant n° 503480), Alzheimer’s Research UK (Grant n° 503176), the Wellcome Trust (Grant n° 082604/2/07/Z) and German Federal Ministry of Education and Research (BMBF): Competence Network Dementia (CND) grant n° 01GI0102, 01GI0711, 01GI0420. CHARGE was partly supported by the NIH/NIA grant R01 AG033193 and the NIA AG081220 and AGES contract N01–AG–12100, the NHLBI grant R01 HL105756, the Icelandic Heart Association, and the Erasmus Medical Center and Erasmus University. ADGC was supported by the NIH/NIA grants: U01 AG032984, U24 AG021886, U01 AG016976, and the Alzheimer’s Association grant ADGC–10–196728. Laura Pulkki-Råback was supported by the Jenny and Antti Wihuri Foundation.

## Contributors

ME and CH designed the study and conducted the statistical analyses. ME wrote the first draft of the manuscript. JL and AM calculated the polygenetic risk score with the help of MP. All authors contributed to the interpretation of the results and critical revision of the manuscript for important intellectual content and approved the final version of the manuscript. The corresponding author attests that all listed authors meet authorship criteria and that no others meeting the criteria have been omitted.

## Ethical approval

Ethical approval for data collection was given by the North-West Multi-centre Research Ethics Committee. Study was carried out in accordance with the Declaration of Helsinki of the World Medical Association. The ethical board of the Finnish Institute for Health and Welfare gave ethical permission to use the genetic data.

## Transparency statement

The lead authors (ME and CH) affirms that the manuscript is an honest, accurate, and transparent account of the study being reported; that no important aspects of the study have been omitted; and that any discrepancies from the study as planned (and, if relevant, registered) have been explained.

## ONLINE SUPPLEMENT

### 1) Additional information of dementia assessment

Incident all-cause dementia was defined using the following ICD-9 and ICD-10 codes:

ICD-9: 290.2, 290.3, 290.4, 291.2, 294.1, 331.0, 331.1, 331.2. 331.5

ICD-10: A81.0, F00, F00.0, F00.1, F00.2, F00.9, F01, F01.0, F01.1, F01.2, F01.3, F01.8, F01.9, F02, F02.0, F02.1, F02.2, F02.3, F02.4, F02.8, F03, F05.1, F10.6, G30, G30.0, G30.1, G30.8, G30.9, G31.0, G31.1, G31.8, I67.3

Incident Alzheimer’s disease was defined using the following ICD-9 and ICD-10 codes:

ICD-9: 331.0

ICD-10: F00, F00.0, F00.1, F00.2, F00.9, G30, G30.0, G30.1, G30.8, G30.9

For more information of the dementia assessment see: http://biobank.ndph.ox.ac.uk/showcase/showcase/docs/alg_outcome_dementia.pdf

### 2) Additional information of genetic risk score

International Genomics of Alzheimer’s Project (IGAP) is a large three-stage study based upon genome-wide association studies (GWAS) on individuals of European ancestry. In stage 1, IGAP used genotyped and imputed data on 11,480,632 single nucleotide polymorphisms (SNPs) to meta-analyse GWAS datasets consisting of 21,982 Alzheimer’s disease cases and 41,944 cognitively normal controls from four consortia: The Alzheimer Disease Genetics Consortium (ADGC); The European Alzheimer’s disease Initiative (EADI); The Cohorts for Heart and Aging Research in Genomic Epidemiology Consortium (CHARGE); and The Genetic and Environmental Risk in AD Consortium Genetic and Environmental Risk in AD/Defining Genetic, Polygenic and Environmental Risk for Alzheimer’s Disease Consortium (GERAD/PERADES). In stage 2, 11,632 SNPs were genotyped and tested for association in an independent set of 8,362 Alzheimer’s disease cases and 10,483 controls. Meta-analysis of variants selected for analysis in stage 3A (n = 11,666) or stage 3B (n = 30,511) samples brought the final sample to 35,274 clinical and autopsy-documented Alzheimer’s disease cases and 59,163 controls.

### 3) The associations between genetic risk score and incident dementia using 10 various geneic risk score cut-off points

**SFigure 1.**
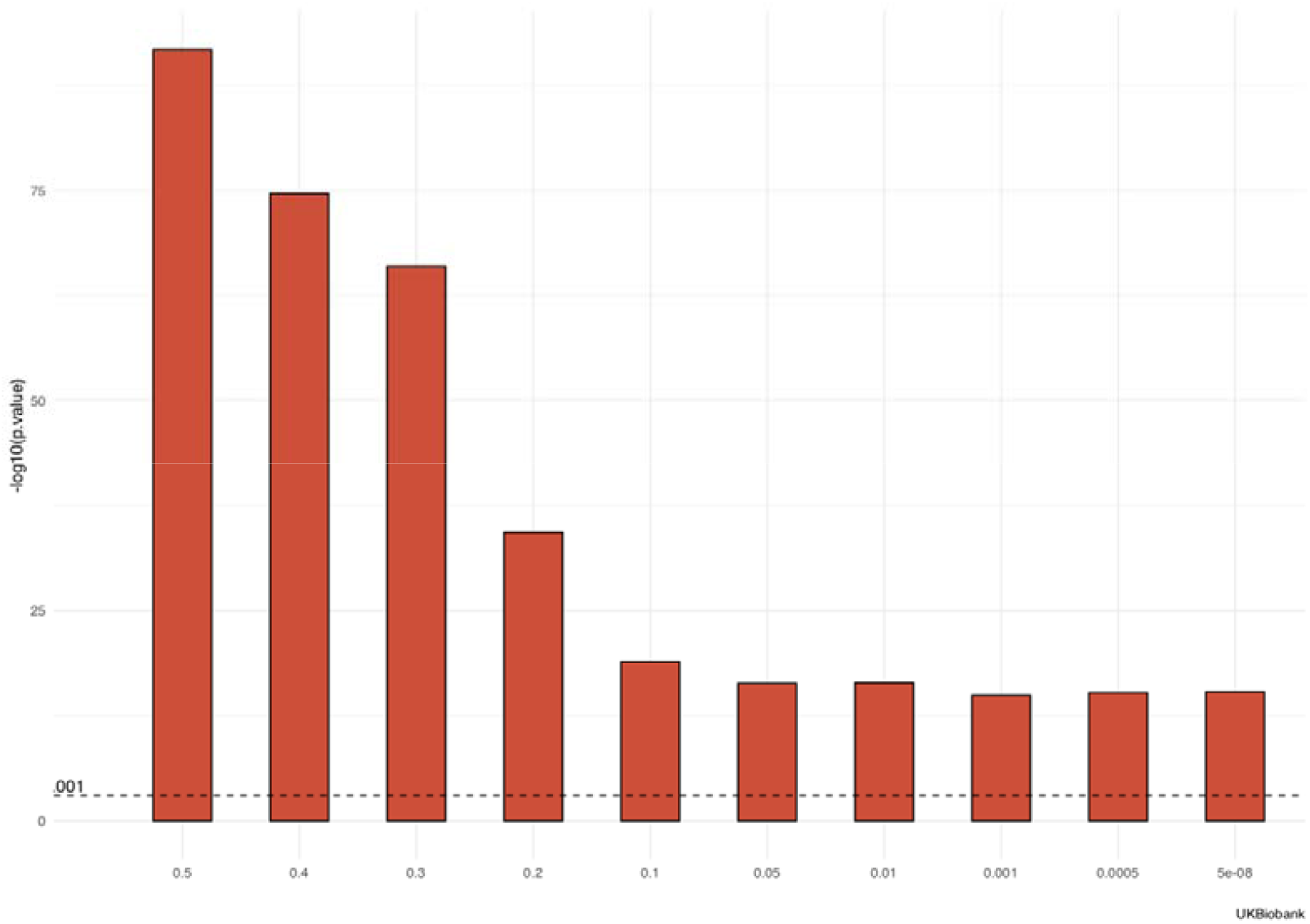
The associations between continuous PRS and incident dementia with various cut-off points. The bars are negative log10 -transformed p-values of the PRS-dementia association.

### 4) The associations of social isolation, loneliness and genetic risk score with specific Alzheimer’s disease

We repeated all the analyses using specific Alzheimer’s disease as the outcome instead of incident dementia and the results were materially the same, although there were, of course, much less Alzheimer’s disease cases.

**STable 2.** Risk of Incident Alzheimers’ Disease According to Genetic Risk

| *Genetic risk* | **Model 1** |  | **Model 2** |  |
| --- | --- | --- | --- | --- |
|  | *HR*<br>*95 % CI* | *P-Value* | *HR*<br>*95 % CI* | *P-Value* |
| Intermediate | 1.91<br>(1.45 – 2.51) | **<0.001** | 1.84<br>(1.35 – 2.50) | **<0.001** |
| High | 2.51<br>(1.86 – 3.38) | **<0.001** | 2.43<br>(1.74 – 3.40) | **<0.001** |
| Observations | 155074 |  | 132628 |  |
| R<sup>2</sup> Nagelkerke | 0.345 |  | 0.392 |  |

Model 1. Adjusted for age, sex, and 10 principal components
Model 2. Adjusted for age, sex, 10 principal components, education, social deprivation, depressive symptoms, health behaviors, loneliness and social isolation

**STable 3.**
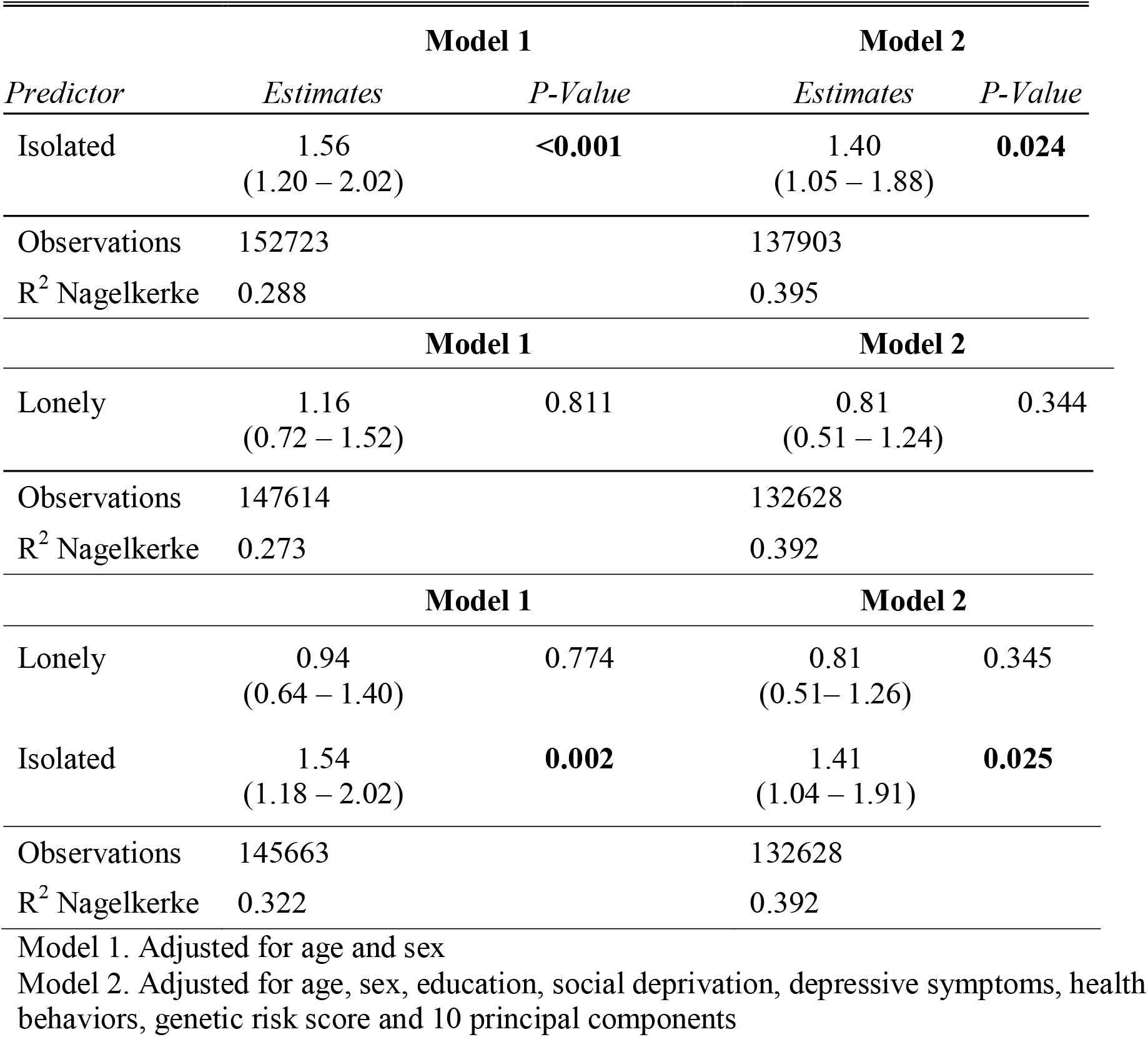
Risk of Incident Azheimers’ Disease According to Social Isolation and Loneliness Categories

| *Predictor* | **Model 1** |  | **Model 2** |  |
| --- | --- | --- | --- | --- |
|  | *Estimates* | *P-Value* | *Estimates* | *P-Value* |
| Isolated | 1.56<br>(1.20 – 2.02) | **<0.001** | 1.40<br>(1.05 – 1.88) | **0.024** |
| Observations | 152723 |  | 137903 |  |
| R<sup>2</sup> Nagelkerke | 0.288 |  | 0.395 |  |
|  | **Model 1** |  | **Model 2** |  |
|  | *Estimates* | *P-Value* | *Estimates* | *P-Value* |
| Lonely | 1.16<br>(0.72 – 1.52) | 0.811 | 0.81<br>(0.51 – 1.24) | 0.344 |
| Observations | 147614 |  | 132628 |  |
| R<sup>2</sup> Nagelkerke | 0.273 |  | 0.392 |  |
|  | **Model 1** |  | **Model 2** |  |
|  | *Estimates* | *P-Value* | *Estimates* | *P-Value* |
| Lonely | 0.94<br>(0.64 – 1.40) | 0.774 | 0.81<br>(0.51– 1.26) | 0.345 |
| Isolated | 1.54<br>(1.18 – 2.02) | **0.002** | 1.41<br>(1.04 – 1.91) | **0.025** |
| Observations | 145663 |  | 132628 |  |
| R<sup>2</sup> Nagelkerke | 0.322 |  | 0.392 |  |

Model 1. Adjusted for age and sex
Model 2. Adjusted for age, sex, education, social deprivation, depressive symptoms, health behaviors, genetic risk score and 10 principal components

**SFigure 2a.**
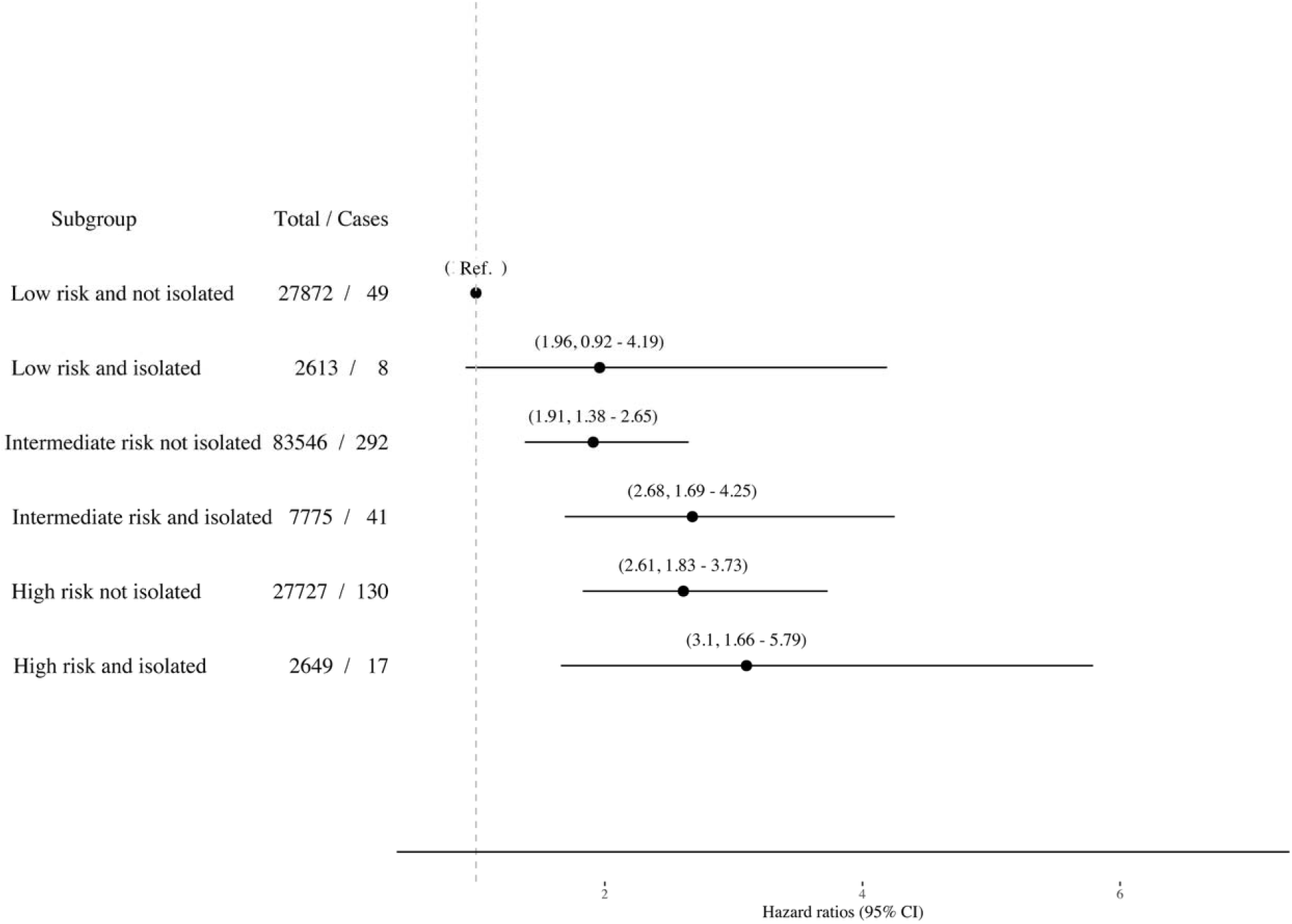
The associations (Hazard ratios and 95% Cis) of combined genetic risk and isolation categories with incident Alzheimer’s disease.

**SFigure 2b.**
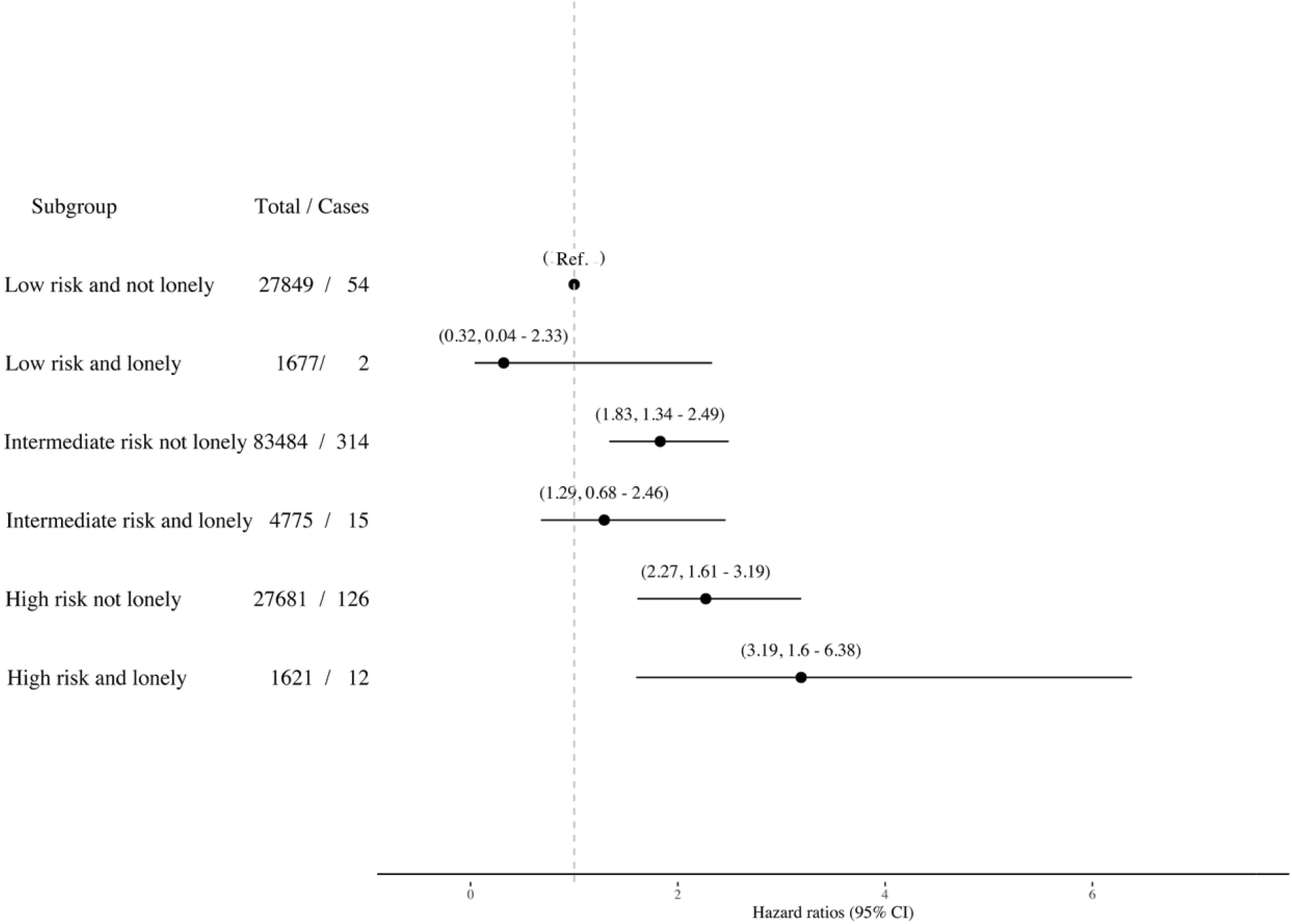
The associations (Hazard ratios and 95% Cis) of combined genetic risk and loneliness categories with incident Alzheimer’s disease.

### 5) The associations of combined social isolation/loneliness and genetic risk score categories with incident dementia using imputed data

The number of missing values was relatively small (only less the 5% had missing values), but we repeated the final models using five imputed data sets and, not surprisingly, the results were materially not changed (SFigure 3).

**SFigure 3.**
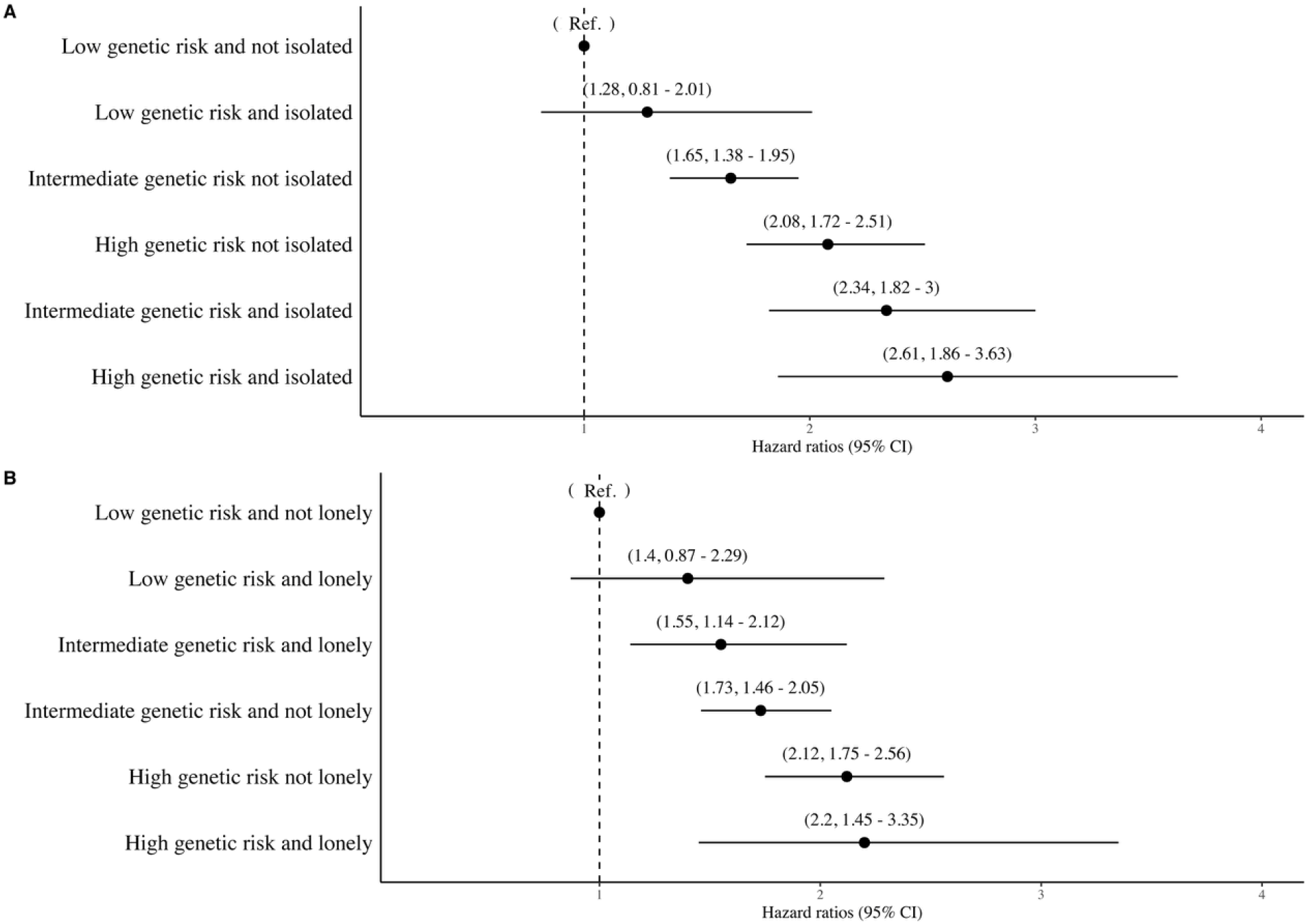
The association (Hazard ratios and 95% Cis) of combined genetic risk and loneliness categories with incident dementia using imputed data (N = 155 070)

### 6) The sex stratified analyses of the final models

We stratified the data according sex and repeated the final analyses using these two data sets. There were only small differences between men and women in any of the associations (sTable 4).

**sTable 4.** Sex stratified associations of combined genetic risk/isolation and genetic risk / loneliness with incident dementia

| **Women ( N= 80 452)** |  |  |  |  |  |
| --- | --- | --- | --- | --- | --- |
| **Group** | **No. of participants** | **Cases** | **HR** | **95% CI** |  |
| Low risk and not isolated | 14550 | 63 | 1.00 | 1.00 | 1.00 |
| Low risk and isolated | 1375 | 8 | 1.14 | 0.52 | 2.50 |
| Intermediate risk not isolated | 43284 | 332 | 1.66 | 1.24 | 2.22 |
| Intermediate risk and isolated | 3966 | 57 | 2.42 | 1.62 | 3.62 |
| High risk not isolated | 14108 | 127 | 1.96 | 1.41 | 2.72 |
| High risk and isolated | 1357 | 19 | 2.35 | 1.30 | 4.24 |
|  | **No. of participants** | **Cases** | **HR** | **95% CI** |  |
| Low risk and not lonely | 14548 | 64 | 1.00 | 1.00 | 1.00 |
| Low risk and lonely | 890 | 7 | 1.27 | 0.51 | 3.18 |
| Intermediate risk not lonely | 43040 | 356 | 1.72 | 1.30 | 2.29 |
| Intermediate risk and lonely | 2608 | 26 | 1.58 | 0.95 | 2.64 |
| High risk not lonely | 14033 | 133 | 1.92 | 1.39 | 2.66 |
| High risk and lonely | 866 | 10 | 2.14 | 1.05 | 4.36 |
| **Men ( N = 74618)** |  |  |  |  |  |
| **Group** | **No. of participants** | **Cases** | **HR** | **95% CI** |  |
| Low risk and not isolated | 13218 | 90 | 1.00 | 1.00 | 1.00 |
| Low risk and isolated | 1225 | 13 | 1.64 | 0.91 | 2.96 |
| Intermediate risk not isolated | 39813 | 410 | 1.51 | 1.18 | 1.93 |
| Intermediate risk and isolated | 3738 | 55 | 1.95 | 1.35 | 2.82 |
| High risk not isolated | 13439 | 186 | 2.00 | 1.52 | 2.63 |
| High risk and isolated | 1267 | 23 | 2.42 | 1.47 | 3.98 |
| **Group** | **No. of participants** | **Cases** | **HR** | **95% CI** |  |
| Low risk and not lonely | 13201 | 90 | 1.00 | 1.00 | 1.00 |
| Low risk and lonely | 771 | 11 | 1.43 | 0.71 | 2.87 |
| Intermediate risk not lonely | 39970 | 433 | 1.57 | 1.23 | 2.00 |

| Women ( N= 80 452) |  |  |  |  |  |
| --- | --- | --- | --- | --- | --- |
| Group | No. of participants | Cases | HR | 95% CI |  |
| Intermediate risk and lonely | 2124 | 32 | 1.42 | 0.89 | 2.27 |
| High risk not lonely | 13466 | 177 | 1.92 | 1.46 | 2.52 |
| High risk and lonely | 742 | 16 | 2.23 | 1.23 | 4.04 |

